# Knowledge, attitudes and practices regarding risk factors for cardiovascular disease among women in an urban slum of the Kathmandu Valley, Nepal: A cross-sectional study

**DOI:** 10.64898/2026.06.04.26354909

**Authors:** Megha Kasaju, Anmol Prasad Shrestha, Natalia Oli, Abhinav Vaidya

## Abstract

**Introduction:** Cardiovascular diseases (CVDs) are the leading cause for death and disability worldwide accounting for 75% of deaths in low- and middle-income countries (LMICs) like Nepal. Urbanization and globalization remains the major cause of rise in CVDs among urban poor population along with growth in slum settlements. This study aims to assess the knowledge, attitude and practice (KAP) of CVDs and its risk factors among women of one such urban poor community in Nepal.

**Methodology:** This cross-sectional study (n=388) in the Sinamangal-Minbhawan slum area was conducted using semi structured questionnaire based on STEPs survey and HARDIC study among the participants selected through convenient sampling. Descriptive analysis was done using SPSS version 21 and KAP scores were further categorized based on median score to perform multivariate logistic analysis. Additionally, Anthropometric and blood pressure measurements were also recorded and analyzed.

**Results:** The median age (Interquartile range) of participants was 33 years (17) with majority of them being Dalit by ethnicity, housewives, with up to primary level education belonging to upper lower socioeconomic class. More than half (53.3%) of the participants were obese and over 23% were hypertensive. While half of the hypertensive women were aware of their status, only 3% had their blood pressure under control. The median knowledge, attitude and practice (KAP) scores were 12, 60 and 10 respectively. The KAP scores were positively associated with socioeconomic status of the participants.

**Conclusion:** The study revealed low knowledge with high prevalence of behavioral risk factors of CVDs along with high prevalence of other metabolic risk factors like high body mass index, high waist hip ratio and hypertension among women of slum area with a positive attitude to prevent CVDs and its risk factors.

## Introduction

Non-communicable diseases (NCDs) are responsible for 70% of global deaths annually.[1] Cardiovascular diseases (CVDs) account for most of NCDs related deaths leading to a worldwide epidemic with cases nearly doubled between 1990 to 2019.[2,3] In Nepal, NCDs caused around 71% deaths in 2019 with 24% of deaths and 12% of DALYs contributed by cardiovascular diseases.[4]

Epidemiological transition is the leading reason for the sudden rise in CVDs.[5] Modern lifestyle changes have resulted in increase in behavioral risk factors like smoking, alcohol consumption, stress, physical inactivity and unhealthy diet as well as metabolic risk factors like obesity, high blood pressure, high blood sugar and cholesterol level [6,7]. Once known as the disease of rich, today the CVD burden is found to be higher among people from low socio-economic group, secluded tribe, low education level with around 75% of the global CVD mortality in low- and middle-income countries (LMICs) due to poor access to affordable health care services for early detection and treatment.[8,9,10]

Moreover, urbanization and globalization have also led to the growth of slum settlements and urban poor population.[11] A slum is a residential area with substandard housing that is poorly serviced and/or overcrowded, and therefore unhealthy, unsafe, and socially undesirable.[12] The urban poor population has reached to 1 billion globally and the burden is increasing in Nepal as well, with around 10 % of population of most cities living in slum area and around 75 slum settlements in Kathmandu valley alone. Living in poor conditions and adapting to a fast-paced urban society makes people living in the slum are vulnerable to various communicable diseases and nutritional deficiencies as-well-as non-communicable diseases.[13,14]

Despite the global pandemic of CVDs, the access to health care system remains differential being limited to urban, rich and male population rendering women from poor socioeconomic region at higher risk of CVDs.[15] Although urban poor women face a substantial disadvantage of poor health literacy, socio-economic deprivation, environmental risk factors, psychosocial abuse with high prevalence of CVDs and its risk factors, this vulnerable group remains neglected with limited research done on the determinants of CVDs among them. Therefore, this study attempts to explore the knowledge, attitude, and practice (KAP) regarding CVD and their risk factors.

## Materials and methods

### Study design and setting

A cross-sectional survey was done to assess the knowledge, attitude, and practice of risk factors for CVDs among women living in Sinamangal-Minbhawan slum of Kathmandu. The data was collected from December 2021 to February 2022.

The Sinamangal-Minbhawan slum area was chosen as it is one of the largest slum areas of Kathmandu valley, and an earlier study revealed a high prevalence of CVD risk factors among male and female population in this area.[16]

### Sampling method and sample size

We selected women of reproductive age (15-49yrs) through convenient sampling as most houses in the area were on rent and the list of the actual slum dwellers were unavailable. The total sample size was calculated as 385 using Cochrane formula (Z^2^pq/e^2^) and keeping prevalence of knowledge regarding risk factors of CVDs at 50% from a previous study.[17] The inclusion criteria was women of reproductive age group (between age 15-49 years) of Sinamangal-Minbhawan slum area who lived there since at least 6 months. One participant was taken from each household of the community.

### Data collection tools and technique

We collected data for assessment of KAP regarding CVDs using a semi-structured questionnaire through one-to-one interview with participants. Knowledge and attitude questions were mostly based on HARDIC trial and other previous CVD studies which were divided into three sections.[17,18,19,20] For the practice questions and physical measurements, we followed the STEPS survey 2019 manual.[21] The first section of the questionnaire was about sociodemographic profile, second section was about knowledge containing questions related to symptoms of CVDs and risk factors of CVDs. The third section contained questions with 5-point Likert scale related to attitude about CVDs and its risk factors and the fourth section was practice section containing questions related to prevalence of various risk factors of CVDs. We also recorded their blood pressure and anthropometric profile (weight, height, waist and hip circumference) during the survey. Weight was measured using a Microlife weighing scale. Height, waist and hip circumference were measured using non-stretchable tapes. Blood pressure was measured manually using a Doctor aneroid sphygmomanometer. Three readings for each respondent were taken at 5 minutes intervals and the average of the last two readings was then recorded.

### Ethical aspects

Ethical clearance was sought from the Institutional Review Committee of Kathmandu Medical College on 11^th^ Aug, 2021 (Ref no. 0608202101). Local leaders of the municipality were informed about the study in detail and their permission was obtained. Written informed consents were taken from all the selected participants. The participants could leave at any time of the interview.

### Data management and analysis

Data was collected through google forms and was analyzed using SPSS version 21.0. The knowledge questions were scored 1 for each correct answer and 0 for each incorrect answer. The attitude questions were scored from 1 to 5 whereas practice questions related to each risk factor was scored 2 for positive behavior, 1 for past users (tobacco, alcohol), and 0 for negative behavior, and practice for health seeking behavior was scored 1. For physical activity, the scores were based on the MET-minutes/week. The maximum possible scores for knowledge, attitude and practice were 38, 75 and 15 respectively. KAP scores were further categorized into two categories for further analysis based on median score obtained as median split is insensitive to extreme values, reduces total deviation and has been proved as a powerful and clear method for analysis.[22,23,24] Body mass index (BMI) of the participants were calculated from their weight and height, and were classified as underweight, normal, overweight, and obese.[25] Similarly, waist-hip ratio (WHR) was classified as increased WHR (>0.85) and normal WHR (≤0.85) based on WHO guidelines.[26] We classified blood pressure measurements as normotension, pre-hypertension, and hypertension according to JNC-8 guidelines.[27] Hypertension care cascade was also assessed. Operational definitions are provided in supplementary file (Text in S1 File). Descriptive analysis including frequency, percentage, median, interquartile range (IQR) and multivariate logistic regression were performed. P-value of <0.05 was considered statistically significant. Multivariate logistic regression was performed among 310 participants only as analysis of socioeconomic status could not be done due to 78 participants refusing to give information about their income.

## Results

### Socio-demographic profile

The median age ± IQR of the participants was 33 ± 17 years with the majority of them (30.4%) belonging to the 26-35 years age group. Most participants were of Dalit ethnicity (43.8%), whereas nearly 70% had primary or lower educational level. Four out of five participants were currently married, majority (82%) belonged to nuclear families, and most (45.4%) were housewives. Only 310 participants could be analyzed for modified Kuppuswamy scale for socioeconomic status as 78 participants refused to give details about their income. Among the 310 participants, two-thirds belonged to upper-lower socio-economic class (Table 1).

**Table 1:** Sociodemographic characteristics of study participants.

| Demographic variables | Frequency (n=388) | Percentage (%) |
| --- | --- | --- |
| **Age (years)** |  |  |
| **≤35** |  |  |
| 15-25 | 102 | 26.3 |
| 26-35 | 118 | 30.4 |
| **>35** |  |  |
| 36-45 | 114 | 29.4 |
| >45 | 54 | 13.9 |
| **Ethnicity** |  |  |
| Dalit | 170 | 43.8 |
| Janajati | 115 | 29.6 |
| Brahmin/chhetri | 78 | 20.1 |
| Muslim | 25 | 6.4 |
| **Education level** |  |  |
| **≤Primary level** |  |  |
| No education | 66 | 17.0 |
| Literate<sup>a</sup>/Primary level<sup>b</sup> | 204 | 52.6 |
| **>Primary level** |  |  |
| Secondary level<sup>c</sup> | 66 | 17.0 |
| More than secondary | 52 | 13.4 |
| **Marital status** |  |  |
| Currently married | 320 | 82.5 |
| **Currently unmarried** |  |  |
| Previously married<sup>d</sup> | 14 | 3.6 |
| Never married | 54 | 13.9 |
| **Family type** |  |  |
| Nuclear | 318 | 82.0 |
| Non- Nuclear<sup>e</sup> | 70 | 18.0 |
| **Occupation** |  |  |
| **Not employed** |  |  |
| Housewife | 176 | 45.4 |
| Student | 27 | 7.0 |
| Unemployed (able to work) | 21 | 5.4 |
| **Employed** |  |  |
| Non formal jobs<sup>f</sup> | 141 | 36.4 |
| Formal jobs<sup>g</sup> | 23 | 5.9 |
| **Socioeconomic status\***<br>**(Modified Kuppuswamy scale)** |  |  |
| Lower class | 2 | 0.5 |
| Lower middle class | 40 | 10.3 |
| Upper lower | 261 | 67.3 |
| Upper middle | 7 | 1.8 |
| Total | 310 | 79.9 |

a: can read and write, b: upto 5<sup>th</sup> grade, c: upto 10<sup>th</sup> grade
d: divorced, separated, widowed, e: joint family, three generation family
f: owns a shop, farmer, labor worker, g: government/non-government jobs
\*78 participants refused to give details about their income

### Metabolic risk factors

Out of the 388 participants, 42% had normal BMI, 34% were overweight, and 19.3% were obese. Similarly, nearly 6 out of 10 participants showed increased WHR (i.e. >0.85). Based on manual measurement of blood pressure, we identified 68 (17.5%) hypertensive and 187 (48.2%) pre-hypertensive participants.

### Hypertension care cascade

We used manual blood pressure measurement along with self-reporting to estimate the actual prevalence of high blood pressure. Among the 388 participants, 92 (23.7%) had hypertension, out of which 46 (50%) were aware about their disease status, 9 (10%) were under treatment and 3(3%) had their blood pressure under control (Fig 1).

**Fig 1:**
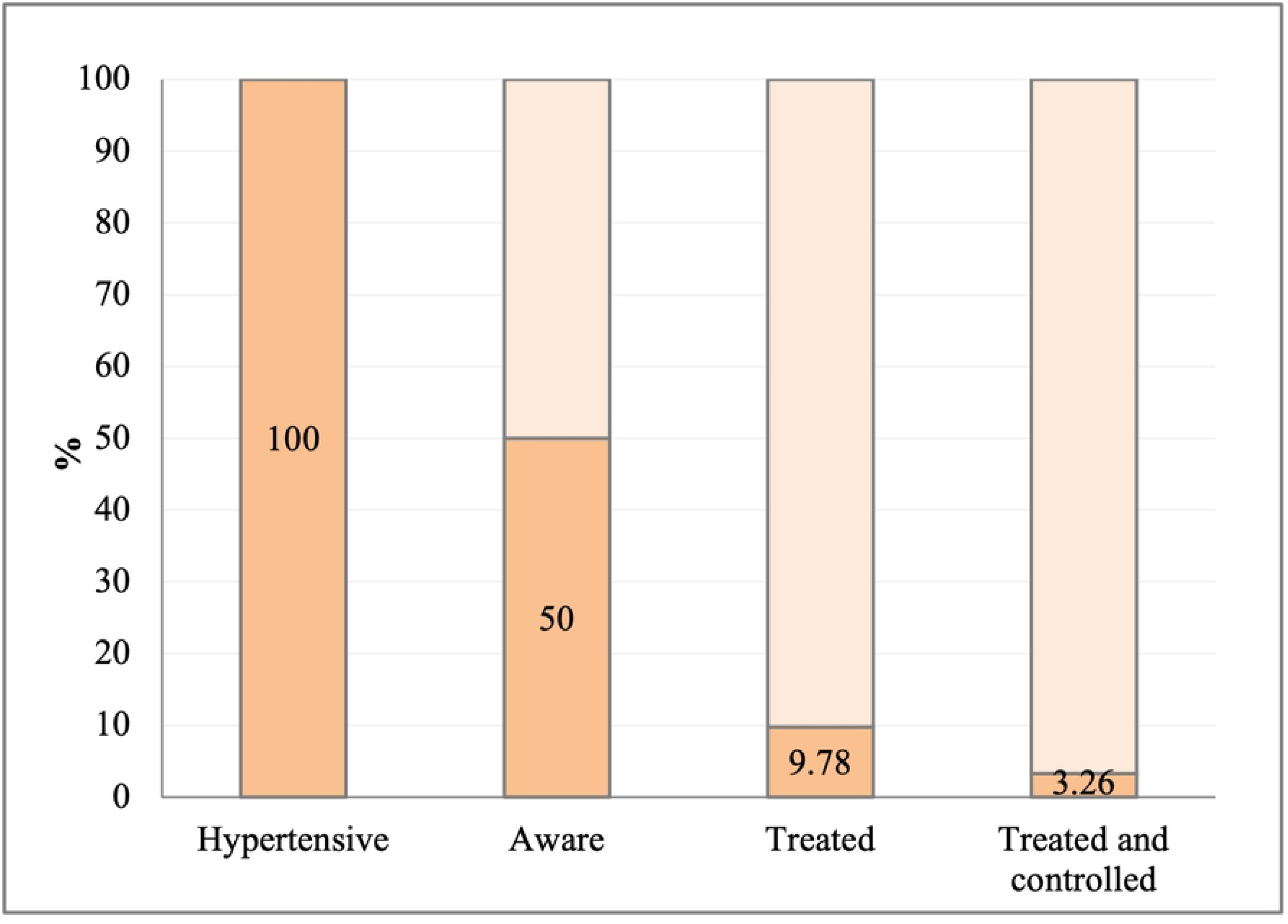
Hypertension care cascade (n = 92)

### Knowledge regarding CVD and its risk factors

We asked the participants to spontaneously list the causes for heart disease which revealed low knowledge about the risk factors among the participants. We again asked the question related to risk factor in a close-ended prompted manner with ‘yes’ and ‘no’ as options, the knowledge level about individual risk factor increased from the non-prompted response (Fig 2). For example, only 4.6% participants could list stress as CVD risk factor during non-prompted response which increased to 41% during prompted response and similar changes were seen for other risk factors. When asked about the symptoms of CVDs, only two-fifth were able to identify chest pain/tightness followed by shortness of breath. Most of them were unaware about other CVD symptoms (Fig 3).

**Fig 2:**
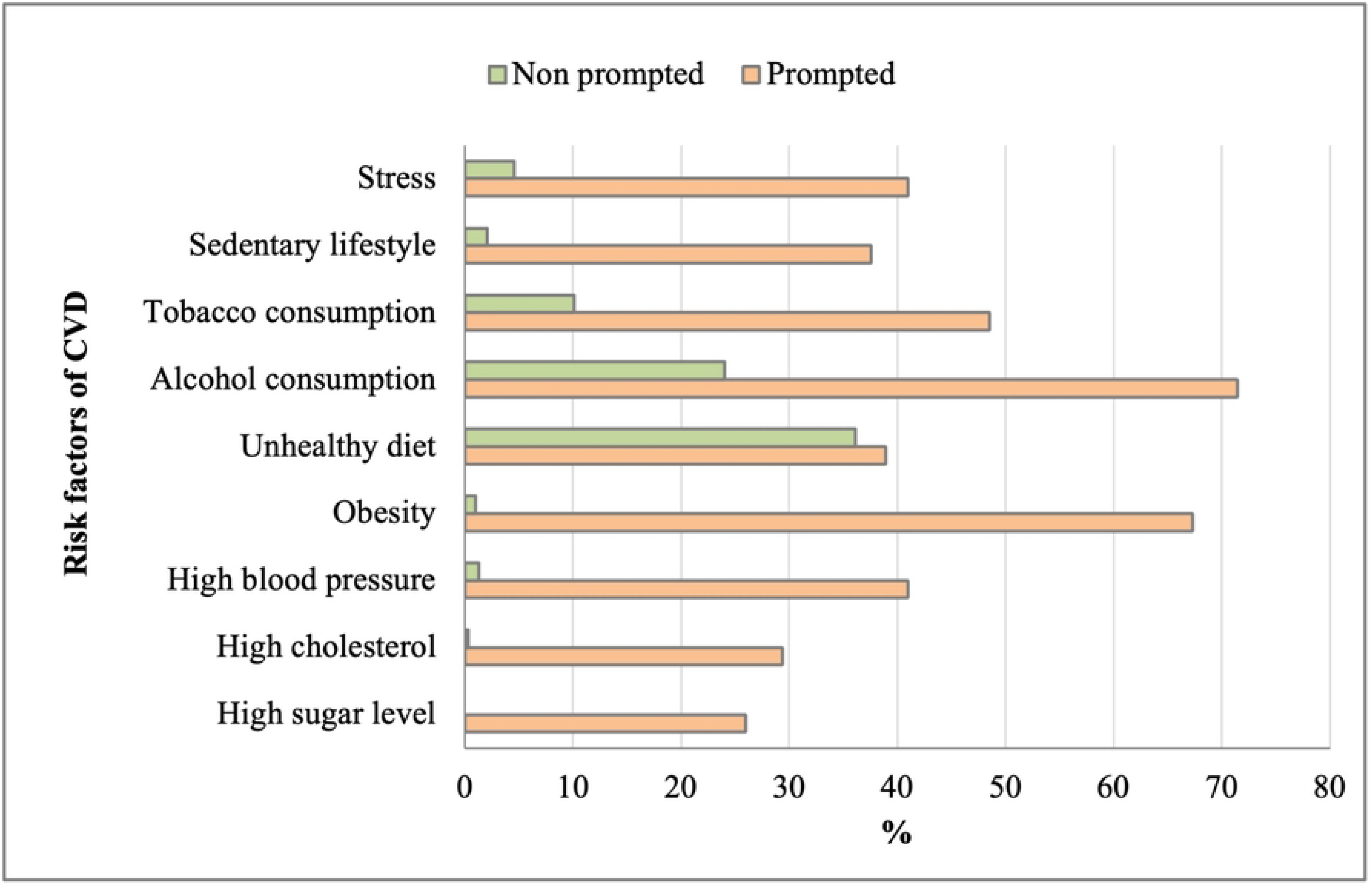
Comparison of prompted and non-prompted responses (%) for risk factors of CVD (multiple responses)

**Fig 3:**
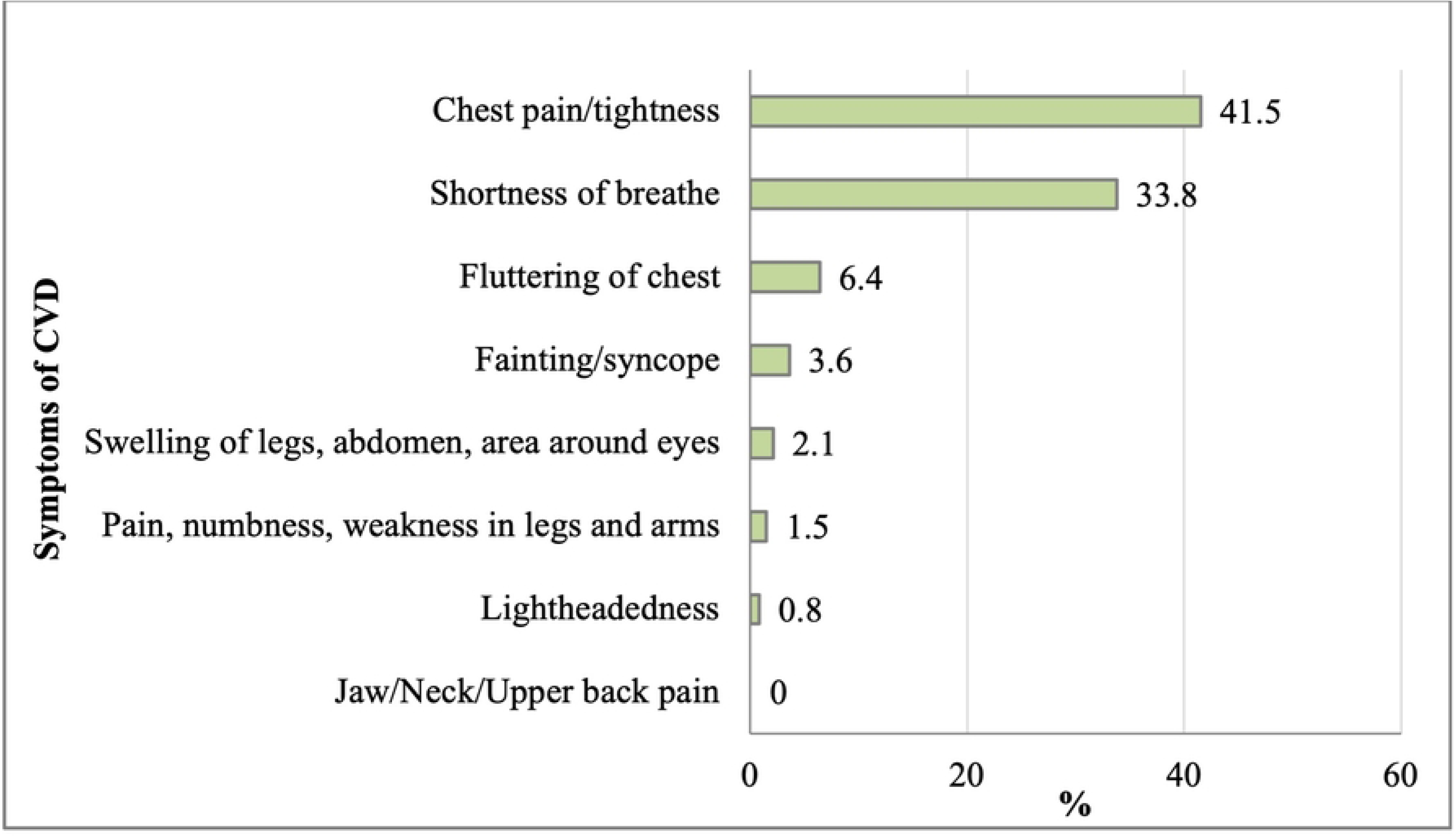
Responses (%) for symptoms of cardiovascular disease (multiple responses)

### Attitude regarding CVD risk factors and their prevention

Most of the participants believed that smoking, stress, unhealthy diet and alcohol had negative effect on their health whereas doing physical activity and eating fruits and vegetables had positive effects. Around half the participants (strongly agree-25.5%, agree-33.8%) were confident about changing their lifestyle. Most of them believed that ready availability of cheaper fruits and vegetables, better built-in environment facilities, better health services, good public policies and awareness program could help them improve their lifestyle (Table 2).

**Table 2:** Responses on attitude related questions on cardiovascular health.

|  | **Strongly Agree** | **Agree** | **Neutral** | **Disagree** | **Strongly disagree** |
| --- | --- | --- | --- | --- | --- |
|  | **N (%)** | **N (%)** | **N (%)** | **N (%)** | **N (%)** |
| **Attitude about risk factors of CVDs** |  |  |  |  |  |
| 1. I think that smoking in small quantities doesn't increase risk of heart disease | 12 (3.1) | 30 (7.7) | 53 (13.7) | 224 (57.7) | 69 (17.8) |
| 2. I think that doing exercise or any physical activity will reduce the risk of heart disease | 116 (29.9) | 229 (59.0) | 36 (9.3) | 1 (0.3) | 6 (1.5) |
| 3. I think eating fruits and vegetables increases risk of heart disease | 8 (2.1) | 12 (3.1) | 62 (16.0) | 184 (47.4) | 122 (31.4) |
| 4. I believe stress management should be done to prevent heart disease | 116 (29.9) | 230 (59.3) | 40 (10.3) | 2 (0.5) | 0 |
| 5. I believe that cold drinks such as coke are healthy | 2 (0.5) | 6 (1.5) | 63 (16.2) | 189 (48.7) | 128 (33) |
| 6. In my opinion, packed fruit juices such as Frooti are as healthy as whole fruit | 12 (3.1) | 13 (3.4) | 13 (3.4) | 224 (57.7) | 126 (32.5) |
| 7. I think heavy alcohol consumption is good for health | 0 | 1 (0.3) | 5 (1.3) | 175 (45.1) | 207 (53.4) |
| **Attitude towards preventive aspects of CVDs** |  |  |  |  |  |
| 1. I believe i can change my lifestyle | 99 (25.5) | 131 (33.8) | 129 (33.2) | 9 (2.3) | 20 (5.2) |
| 2. In my opinion, healthy foods including fruits and vegetables are costlier, hence quite unaffordable | 43 (11.1) | 220 (56.7) | 57 (14.7) | 32 (8.2) | 36 (9.3) |
| 3. I think greater access to public recreational activities can improve physical activity | 121 (31.2) | 202 (52.1) | 51 (13.1) | 7 (1.8) | 7 (1.8) |
| 4. In my opinion, tobacco consumption should be banned | 250(64.4) | 106(27.3) | 23(5.9) | 7(1.8) | 2(0.5) |
| 5. I believe there should be more awareness program for healthy heart | 184(47.4) | 198(51.0) | 5(1.3) | 1(0.3) | 0 |
| 6. I think there should be availability of health facility for heart disease irrespective of socioeconomic status | 178(45.9) | 205(52.8) | 5(1.3) | 0 | 0 |

### Practice regarding CVD risk factors

Around half of the participants (50.8%) were under stress and almost all the participants (98.2%) had low fruit and vegetable intake. Nearly 10% had inadequate physical activity whereas around 12% were current alcohol consumers and 16% were current smokers (Fig 4).

**Fig 4:**
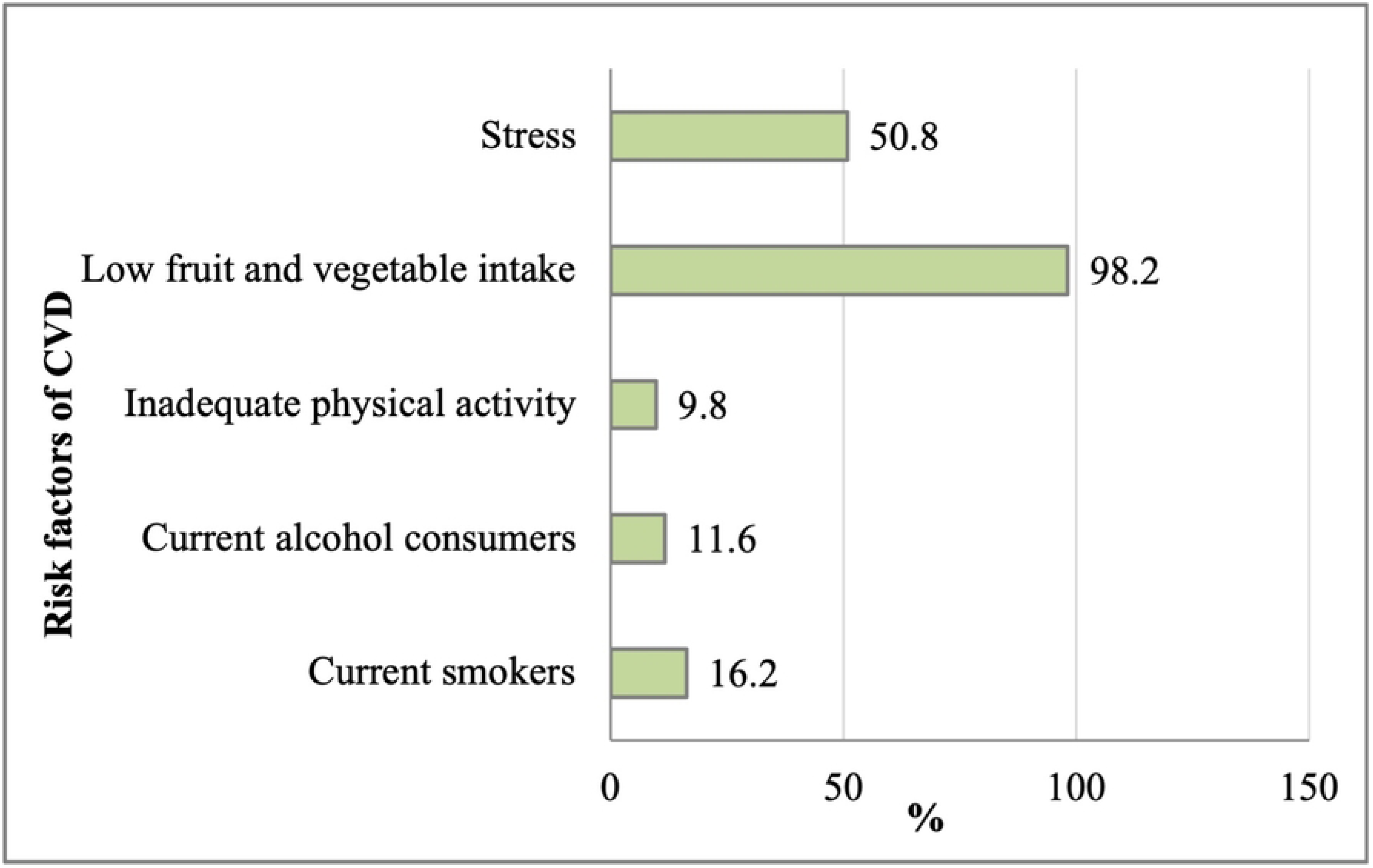
Prevalence of behavioral risk factors in Sinamangal-Minbhawan slum area

### Knowledge, Attitude, and Practice scores

The median KAP scores were 12, 60 and 10 respectively. The KAP scores were further categorized based on the median scores to find the association with other independent variables (Table 3).

**Table 3:** Knowledge, attitude, and practice score (categorized based on median)

| | Median (IQR) | Below median<br>($\leq 50^{\text{th}}$ percentile) | Above median<br>($>50^{\text{th}}$ percentile) |
| --- | --- | --- | --- |
| Knowledge | 12 (6) | 235(60.6) | 153(39.4) |
| Attitude | 60 (7) | 198(51.0) | 190 (49.0) |
| Practice | 10 (2) | 247 (63.7) | 141 (36.3) |

SD: Standard deviation, IQR: Interquartile range

### Association of KAP with various independent variables

After adjusting for other confounders, knowledge was significantly associated with socio-economic status (AOR = 7.57, 95% CI: 3.59-15.98) (Table 4). Similarly, attitude was associated with family type (AOR = 7.57, 95% CI: 3.59-15.98) and socio-economics status (AOR = 5.19, 95% CI: 2.41-11.19) (Table 5). Practice was associated with age (AOR = 0.53, 95% CI: 0.32-0.89) and socio-economic status (AOR = 3.43, 95% CI: 1.75-6.70), after adjusting for other independent variables (Table 6).

**Table 4:** Relationship of knowledge to sociodemographic variables.

| **Sociodemographic profile** | **Above median**<br>**N (%)** | **Below median**<br>**N (%)** | **COR (95% CI)**<br>***p value*** | **AOR (95% CI)**<br>***p value*** |
| --- | --- | --- | --- | --- |
| **Age (years)** |  |  |  |  |
| >35 | 58 (34.5) | 110 (65.5) | 0.694 (0.458-1.051)<br>*0.084* | 0.937 (0.560-1.568)<br>*0.803* |
| ≤35 | 95 (43.2) | 125 (56.8) | RC |  |
| **Education level** |  |  |  |  |
| >Primary level | 57 (48.3) | 61 (51.7) | 1.694 (1.092-2.626)<br>*0.019\** | 1.104 (0.624-1.954)<br>*0.734* |
| ≤Primary level | 96 (35.6) | 174 (64.4) | RC |  |
| **Ethnicity** |  |  |  |  |
| Dalit/Janajati/Muslim | 121 (39.0) | 189 (61.0) | 0.920 (0.555-1.526)<br>*0.747* | 1.131 (0.647-1.975)<br>*0.666* |
| Brahmin/Chhetri | 32 (41.0) | 46 (59.0) | RC |  |
| **Marital status** |  |  |  |  |
| Currently married | 116 (36.3) | 204 (63.7) | 0.476 (0.281-0.808)<br>*0.006\** | 0.568 (0.304-1.063)<br>*0.077* |
| Single | 37 (54.4) | 31 (45.6) | RC |  |
| **Occupation** |  |  |  |  |
| Employed | 63 (38.4) | 101 (61.6) | 0.929 (0.615-1.403)<br>*0.725* | 1.033 (0.651-1.637)<br>*0.892* |
| Not employed | 90 (40.2) | 134 (59.8) | RC |  |
| **Family type** |  |  |  |  |
| Nuclear | 117 (36.8) | 201 (63.2) | 0.550 (0.326-0.926)<br>*0.024* | 0.677 (0.386-1.189)<br>*0.175* |
| Non-nuclear | 36 (51.4) | 34 (48.6) | RC |  |

| **Socioeconomic status** |  |  |  |  |
| --- | --- | --- | --- | --- |
| Kuppuswamy score >10 | 36 (76.6) | 11 (23.4) | 7.762 (3.759-16.030)<br><0.01* | 7.572 (3.588-15.981)<br><0.01* |
| Kuppuswamy score ≤10 | 78 (29.7) | 185 (70.3) | RC |  |

COR: Crude odds ratio, AOR: Adjusted odds ratio RC: Reference category, CI: Confidence interval,
\*p-value <0.05

**Table 5:** Relationship of attitude to sociodemographic variables.

| Sociodemographic profile | Above median<br>N (%) | Below median<br>N (%) | COR (95% CI)<br>*p-value* | AOR (95%CI)<br>*p-value* |
| --- | --- | --- | --- | --- |
| Age (years) |  |  |  |  |
| >35 | 78 (46.4) | 90 (53.6) | 0.836 (0.559-1.250)<br>0.382 | 0.900 (0.551-1.470)<br>0.674 |
| ≤35 | 112 (50.9) | 108 (49.1) | RC |  |
| Education level |  |  |  |  |
| >Primary level | 61 (51.7) | 57 (48.3) | 1.170 (0.759-1.803)<br>0.478 | 0.857 (0.491-1.495)<br>0.586 |
| ≤Primary level | 129 (47.8) | 141 (52.2) | RC |  |
| Ethnicity |  |  |  |  |
| Dalit/Janajati/Muslim | 149 (48.1) | 161 (51.9) | 0.835 (0.508-1.373)<br>0.478 | 0.987 (0.581-1.677)<br>0.962 |
| Brahmin/Chhetri | 41 (52.6) | 37 (47.4) | RC |  |
| Marital status |  |  |  |  |
| Currently married | 156 (48.8) | 164 (51.2) | 0.951 (0.564-1.606)<br>0.851 | 1.189 (0.632-2.235)<br>0.591 |
| Currently unmarried | 34 (50.0) | 34 (50.0) | RC |  |
| Occupation |  |  |  |  |
| Employed | 73 (44.5) | 91 (55.5) | 0.734 (0.490-1.099)<br>0.133 | 0.768 (0.493-1.196)<br>0.243 |
| Not employed | 117 (52.2) | 107 (47.8) | RC |  |
| Family type |  |  |  |  |
| Nuclear | 176 (55.3) | 142 (44.7) | 0.370 (0.213-0.642)<br><0.01* | 0.410 (0.231-0.728)<br>0.002* |
| Non-nuclear | 48 (68.6) | 22 (31.4) | RC |  |

| **Socioeconomic status** |  |  |  |  |
| --- | --- | --- | --- | --- |
| Kuppuswamy score >10 | 37 (78.7) | 10 (21.3) | 5.310 (2.532-11.135)<br><0.01* | 5.188 (2.406-11.186)<br><0.01* |
| Kuppuswamy score ≤10 | 108 (41.1) | 155 (58.9) | RC |  |

COR: Crude odds ratio, AOR: Adjusted odds ratio RC: Reference category, CI: Confidence interval
\*p-value <0.05

**Table 6:** Relationship of practice to sociodemographic variables.

| Sociodemographic profile | Above median<br>N (%) | Below median<br>N (%) | COR (95% CI)<br>*p-value* | AOR (95%CI)<br>*p-value* |
| --- | --- | --- | --- | --- |
| Age (years) |  |  |  |  |
| >35 | 45 (26.8) | 123 (73.2) | 0.473 (0.306-0.729)<br>0.001* | 0.534 (0.319-0.894)<br>0.017* |
| ≤35 | 96 (43.6) | 124 (56.4) | RC |  |
| Education level |  |  |  |  |
| >Primary level | 53 (44.9) | 65 (55.1) | 1.686 (1.083-2.627)<br>0.021* | 0.966 (0.551-1.694)<br>0.903 |
| ≤Primary level | 88 (32.6) | 182 (67.4) | RC |  |
| Ethnicity |  |  |  |  |
| Dalit/Janajati/Muslim | 107 (34.5) | 203 (65.5) | 0.682 (0.412-1.130)<br>0.138 | 0.759 (0.443-1.300)<br>0.315 |
| Brahmin/Chhetri | 34 (43.6) | 44 (56.4) | RC |  |
| Marital status |  |  |  |  |
| Currently married | 112 (35.0) | 208 (65.0) | 0.724 (0.425-1.234)<br>0.235 | 1.164 (0.614-2.208)<br>0.903 |
| Currently unmarried | 29 (42.6) | 39 (57.4) | RC |  |
| Occupation |  |  |  |  |
| Employed | 47 (28.7) | 117 (71.3) | 0.556 (0.361-0.854)<br>0.007* | 0.649 (0.407-1.035)<br>0.069 |
| Not employed | 94 (42.0) | 130 (58.0) | RC |  |
| Family type |  |  |  |  |
| Nuclear | 109 (34.3) | 209 (65.7) | 0.619 (0.367-1.046)<br>0.073 | 0.735 (0.422-1.280)<br>0.276 |
| Non-nuclear | 32 (45.7) | 38 (54.3) | RC |  |
| **Socio-economic status** |  |  |  |  |
| Kuppuswamy score >10 | 28 (59.6) | 19 (40.4) | 3.764 (1.982-7.149)<br><0.01* | 3.423 (1.749-6.700)<br><0.01* |
| Kuppuswamy score ≤10 | 74 (28.1) | 189 (71.9) | RC |  |

COR: Crude odds ratio, AOR: Adjusted odds ratio, RC: Reference category, CI: Confidence interval
\*p-value <0.05

## Discussion

The study reports low knowledge, positive attitude, and high prevalence of cardiovascular risk factors among women in the Sinamangal-Minbhawan slum area.

### Socio-demographic profile of slum population

This study has diverse group of participants but most of them have disadvantaged background. Similar findings could be found in a study done in Amritsar, India as well as other slums of Kathmandu valley where most of the inhabitants were of secluded caste with low education level, poor socioeconomic condition mostly involved in manual work.[16,28,29] Most women being involved in employment (working 10-12 hours per day) made it difficult for us to reach our target sample which caused most of our participants to be housewives as they were the reachable population there during time of data collection. This was a similar scenario in other studies as well.[28,30]

### Low knowledge about cardiovascular health

We evaluated prompted and non-prompted responses to see the knowledge of risk factors which is done in a similar study in Nepal, which showed better knowledge of risk factors with prompted questions than non-prompted. The most common symptoms identified by the participants were chest pain and shortness of breath which was also found in a study done in urbanizing community of Nepal.[31] However, another study done in Pakistan identified chest pain and pain in jaws and arms as the most known symptom among participants.[32] The difference in symptom identification between these participants can be because of symptom variation and difference in awareness programs between countries.

### Community perspective about adapting healthy lifestyle

In contrary to the knowledge, attitude of the participants was by and large positive. As in another study, given their current circumstances, with the available limited resources, the lifestyle they follow was all they could follow to sustain their life in the costly urban life.[33] They thought adapting to healthier lifestyle would be unchallenging if there were stricter policies for alcohol and tobacco products, easy availability of healthier food options and better physical activity options. Likewise, a study done in Pakhribas municipality of Eastern Nepal revealed participants willingness to seek health care and change in lifestyle only after diagnosis with CVDs.[34] This study found that barriers to good heart health in urban poor population can be financial difficulties, lack of physical infrastructures, lack of unavailability of healthy food options and health services which was supported by other studies as well.[35,36]

### Higher prevalence of CVD risk factors among slum population

The prevalence of risk factors like smoking, alcohol consumption, low fruit and vegetable intake, physical inactivity in the Sinamangal-Minbhawan slum was higher in the women residing in slum area as compared to general women population whereas stress was lower compared to general population.[21] A qualitative study done in Bangladesh has shown increase in harmful risk factors because of adaptation to urban lifestyle. Lack of healthy food and stress due to poverty, increased consumption of alcohol, tobacco to handle stress, intake of outside food, beverages were seen as some of the major lifestyle issues.[37]

### Obesity and hypertension burden among slum women

More than half of the participants had BMI above 25kg/m^2^ and the finding were similar to a study done in slum dwellers of Tamil Nadu, India.[38] However, in the STEPS survey 2019, only a quarter of the female participants had BMI >25kg/m^2^.[21] The waist hip ratio >0.85 was found in equal percentage in both this survey and STEPS survey 2019 while it was lower in women of slums of northern India.[21,39]

High blood pressure prevalence was found to be like the average Nepali population whereas the awareness status was much higher than that of average Nepali population but very few participants were under treatment and had controlled blood pressure.[21] This may be due to difficulty in access to health center, financial difficulties and non-adherence to medication.

A positive significant relation between BMI and systolic blood pressure (r=0.267, p<0.01) and BMI and diastolic blood pressure (r=0.212, p<0.01) was seen in this study which is also seen in a study done among Chinese population and is supported by another study done in Italy.[40,41] Similarly, a positive relation was also seen between waist-hip ratio and both systolic (r=0.280, p<0.01) and diastolic blood pressure (r=0.326, p<0.01) which is also supported by other studies.[42,43]

### Barriers to accessing primary health care services

In a low-income country like Nepal, there is a disparity in availability of health services in rural and urban settings throughout Nepal.[44] Financial burden, government corruption, poor infrastructure and lack of awareness have resulted in inequality in health care service delivery in Nepal.[45] Although our study area is in the center of capital city with nearest primary healthcare center having PEN package services at distance of 2-3kms and a tertiary care hospital at distance of mere 500m, very few people visit these places to utilize the services.[46] From this study we found that, people still are unaware about the free services provided by Government of Nepal for healthy heart.[47] This finding is similar to a study done in slum of Iran where the main barrier to health service utilization was limited access and lack of awareness among inhabitants.[48]

### Strengths and Limitations

This study is the first cross sectional study on cardiovascular health risk factors to be done among women residing in slum area of Kathmandu valley. From this study alone, we found evidence of poor knowledge, attitude, practice of behavioral risk factors of CVDs and its preventive aspects among urban poor population We also explored the difficulties in day-to-day life of urban poor population and their barriers to adapting a healthy lifestyle and access to health care system which might help the authorities to formulate better policies. This study contributes to the literature of the much unexplored urban poor population of Nepal.

This study was conducted in only one slum of Kathmandu valley therefore the findings may not be generalizable to the other settings. The methodological issues might have created some biases in the study. Moreover, the data on behavioral risk factor practices like physical activity and stress were subjective. Also, the hypertension prevalence included self-reported data, because of which there may be some over estimation or under estimation of hypertension prevalence.

## Conclusion

This study has highlighted low knowledge with high prevalence of harmful risk factors of CVDs among urban poor women despite a positive attitude to prevent CVDs and its risk factors. Major proportion of participants were found to be over-weight and obese with high waist hip ratio. Similarly, the prevalence of high blood pressure was also high among the participants, only half were aware about their disease status whereas very few were under treatment and had their blood pressure under control. These findings reflects a need for sustainable community-based interventions focusing on risk factors of cardiovascular diseases to generate awareness among the underprivileged at-risk population of LMICs like Nepal.

## Data Availability

The data underlying this study cannot be made publicly available due to the potential identification of the women respondents residing in the slum regions within the study sites. The anonymized dataset required to replicate the study findings including the KAP scores and metabolic measurements will be provided upon reasonable request to the corresponding author (MK).

## Acknowledgement

We are grateful to the women of the slum area for providing their time despite their busy daily schedule and participating in the study. We acknowledge the support of the local leaders in completion of the study.

## Supporting Information List

S1 File: Appendix - Operational definitions

## Notes

### Competing Interest Statement

The authors have declared no competing interest.

### Funding Statement

The author(s) received no specific funding for this work.

### Author Declarations

Ethical clearance was sought from the Institutional Review Committee of Kathmandu Medical College on 11th Aug, 2021 (Ref no. 0608202101).

